# Infant and Neonatal Mortality During the Covid-19 Pandemic: An Interrupted Time Series Analysis From Five Low- and Middle-Income Countries

**DOI:** 10.1101/2023.08.03.23293619

**Authors:** Zachary Wagner, Sam Heft-Neal, Zetianyu Wang, Renzhi Jing, Eran Bendavid

## Abstract

**Background:** The Covid-19 pandemic led to widespread changes to health and social institutions. The effects of the pandemic on neonatal and infant health outcomes in low- and middle-income countries (LMICs) are poorly understood, and nationally representative data characterizing changes to health care and outcomes is only now emerging.

**Methods:** We used nationally representative survey data with vital status and perinatal care information on 2,959,203 children born in India, Madagascar, Cambodia, Nepal, and the Philippines. Using interrupted time series models, we estimated the change in neonatal mortality (death in first 30 days of life) and infant mortality (death in first year of life) following the start of the Covid-19 pandemic, controlling for granular location fixed-effects and seasonality.

**Findings:** We analyzed 2,935,052 births (146,820 deaths) before March 2020 and 24,151 births (799 deaths) after March 2020. We estimated that infant mortality increased by 9.9 deaths per 1,000 live births after March 2020 (95% CI 5.0, 15.0; p<0.01; 22% increase) and neonatal mortality increased by 6.7 deaths per 1,000 live births (95% CI 2.4, 11.1; p<0.01; 27% increase). We observe increased mortality in all study countries. We also estimated a 3.8 percentage point reduction in antenatal care use (95% CI -4.9, -2.7; p<0.01) and a 5.6 percentage point reduction in facility deliveries (95% CI -7.2, -4.0; p<0.01) during the pandemic.

**Interpretation:** Since the start of the Covid-19 pandemic, neonatal and infant mortality are higher than expected in five LMICs. Helping LMICs resume pre-pandemic declines in neonatal and infant mortality should be a major global priority.

**Funding:** National Institute of Child Health and Development (R01HD104835 PI Wagner)

**Research in context:** *Evidence before this study:* The impact of the Covid-19 pandemic on infant and neonatal mortality in low- and middle-income countries (LMICs) is not well-understood. We searched PubMed using the terms “COVID” AND ((“child” OR “infant” OR “neonatal”) AND “mortality”)) AND (“low- and middle-income countries” OR “developing countries”) on May 10, 2023, without language restrictions. The existing evidence is mixed. Increased mortality rates have been documented in Ghana, Nigeria, Uganda, and Nepal while decreased rates documented in South Africa and Guinea. Prior analyses were mainly based on clinic and hospital administrative data and were often confined to a selection of facilities or geographic areas, hampering the generalizability of the existing evidence. We found no published article that leveraged nationally representative data sources to provide a general assessment of infant or neonatal mortality in LMICs following the start of the Covid-19 pandemic.

*Added value of this study:* To our knowledge, this study provides the most comprehensive and generalizable investigation of the impact of the Covid-19 pandemic on infant and neonatal mortality in LMICs to date. Using nationally representative survey data from five LMICs that were recently released, we estimated an increase of 9.9 and 6.7 deaths per 1,000 live births in infant and neonatal mortality, respectively, during the Covid-19 pandemic. We also found significant reductions in antenatal care use and facility deliveries, which could partly explain the changes in mortality we document.

*Implications of the available evidence:* Our study highlights significant increases in infant and neonatal mortality rates in five LMICs following the start of the Covid-19 pandemic, which sets back about a decade’s worth of progress. The decline in antenatal care services and facility births documented in our study suggests mortality increases were partly driven by disruptions in health service access induced by Covid-19 control measures. Helping to get reductions in neonatal and infant mortality back on track in LMICs should be a major global priority.

## Introduction

The Covid-19 pandemic led to abrupt changes to social structures around the world, including health care and public health. The health of young children, particularly in low- and middle-income countries (LMICs), could be vulnerable to Covid-19 infections, as well as to disruptions to health care access, reduced vaccination rates, economic hardship and food insecurity.^1-3^ However, pandemic-related changes in infant and neonatal mortality in LMICs are poorly characterized, and evidence has been mixed with some papers finding increased^4-6^ and some finding decreased mortality^7-9^. Moreover, the available evidence is mainly based on clinic and hospital administrative data and often confined to a selection of facilities or geographic areas, hampering generalizability.^4-14^

Over the past few months, nationally-representative health surveys conducted after the pandemic onset have been released. Here, we analyze pandemic-related changes in infant and neonatal mortality, antenatal care utilization, and facility deliveries using data from five LMICs: India, Cambodia, Madagascar, Nepal, and the Philippines. This study provides the most wide-scoping and generalizable evidence to-date on mortality trends among young children during the Covid-19 pandemic.

## Methods

We used an interrupted time series analyses to estimate the changes in neonatal mortality (death in the first month of life) and infant mortality (death before age one) during the pandemic. The analytic approach compares the observed mortality rates to the rates expected based on historical time trends in each country. We also estimated changes in receipt of antenatal care and facility births.

### Data and Outcomes

We used Demographic and Health Survey data to measure mortality. Each survey includes information about all births among a nationally representative sample of women between 15 and 49 years old. This information includes month of birth, vital status, and, for children that died, the age at death. These data are a primary source of national vital statistics in many LMICs. As of July 1, 2023, India, Cambodia, Madagascar, Nepal, and the Philippines have released survey data containing pandemic-era mortality. In addition to the 5 pandemic-era surveys, we included 13 pre-pandemic survey rounds across the five countries to better estimate baseline trends in mortality. We coded a birth as “exposed” to the pandemic if it occurred at any point after March 2020. Births between April 2019 and March 2020 may have partially overlapped with pandemic control measures. In our primary analysis, we coded these births as “unexposed”, and in sensitivity analyses we dropped them from the sample or coded them as exposed.

### Outcomes

Our primary outcomes are neonatal mortality and infant mortality, and our secondary outcomes are any antenatal care use during pregnancy and whether a birth was delivered in a health facility. Both secondary outcomes have been shown to have important effects on child survival.^15-17^

### Statistical Analysis

Our interrupted time series models estimate how observed mortality rates among births that occurred after March 2020 deviated from what would be expected had the pre-Covid-19 trend in mortality continued. Equation 1 portrays our interrupted time series model.

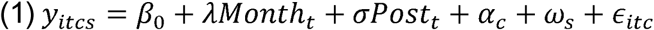

Where *y* is one of our outcomes described above for child *i* born in month *t* in cluster location *c* (roughly village or neighborhood), in month of year *s*. We estimated the pre-Covid-19 trend in mortality at the month level which is represented by *λ*. The term *Post* indicates whether the child was born after March 2020 and the coefficient *σ* is our main estimate of interest, which can be interpreted as the difference between the predicted mortality rate had the pre-Covid-19 trend continued and the observed mortality rate. We included cluster location fixed-effects, *α*_*c*_, to control for time-invariant differences in mortality rates across locations and calendar-month fixed-effects, *ω*_*s*_, to control for seasonality. The underlying assumption of this approach for estimates to be interpreted as causal is that the pre-Covid-19 pandemic linear trend in infant and neo-natal mortality would have continued in absence of the pandemic. In our primary analysis we used all births starting in 1990; we tested the sensitivity of our findings to pre-pandemic trend duration by using shorter historical time windows, up to 2010 as the start year. We applied weights using a combination of the DHS sampling weights and the country population size.^18^ We also estimated results without weights in sensitivity analysis.

## Results

Our sample included 2,935,052 births, 146,021 infant deaths, and 91,113 neonatal deaths before the start of the Covid-19 pandemic, and 24,151 births, 799 infant deaths and 600 neonatal deaths after the start the pandemic (Table 1). The average mother’s age at the time of survey was 36, at the time of birth was 24, and 59% of mothers completed primary education.

**Table 1.** Description of Sample.

|  | Pooled | India | Cambodia | Madagascar | Nepal | Philippines |
| --- | --- | --- | --- | --- | --- | --- |
| Number of births | 2,959,203 | 2,491,193 | 146,755 | 97,649 | 110,479 | 113,127 |
| Pre-March 2020 | 2,935,052 | 2,478,292 | 143,989 | 94,618 | 108,432 | 109,721 |
| Post-March 2020 | 24,151 | 12,901 | 2,766 | 3,031 | 2,047 | 3,406 |
| Number of infant deaths | 146,820 | 120,676 | 10,227 | 5,894 | 7,282 | 2,741 |
| Pre-March 2020 | 146,021 | 120,165 | 10,192 | 5,757 | 7,229 | 2,678 |
| Post-March 2020 | 799 | 511 | 35 | 137 | 53 | 63 |
| Number of neonatal deaths | 91,713 | 78,577 | 4,383 | 2,700 | 4,499 | 1,554 |
| Pre-March 2020 | 91,113 | 78,172 | 4,362 | 2,612 | 4,456 | 1,511 |
| Post-March 2020 | 600 | 405 | 21 | 88 | 43 | 43 |
| Mother's age at survey (years) | 36.1 | 36.3 | 36 | 34.1 | 33.5 | 37.6 |
| Mother's age at birth (years) | 24.2 | 23.9 | 26.7 | 25.1 | 23.1 | 26.4 |
| Mother completed primary school | 59.60% | 57.50% | 73.30% | 73.00% | 38.40% | 97.50% |
| Survey Years |  | 2015-16,<br>2019-22 | 2000,<br>2005,<br>2010,<br>2014,<br>2021 | 1997,<br>2008,<br>2021 | 2001,<br>2006,<br>2011,<br>2016,<br>2022 | 2008,<br>2017,<br>2022 |
| Latest month of birth record | June, 2022 | Apr, 2021 | Dec, 2021 | Jul, 2021 | June, 2022 | June, 2022 |

Data are from demographic and health surveys.

Figure 1 shows infant (panel A) and neonatal (panel B) mortality rates from 2000-2022. The figure shows a steady downward trend in infant and neonatal mortality prior to the start of the pandemic. During the pandemic, both infant and neonatal mortality were higher than expected had this trend continued into the Covid-19 era (pooling years 2020, 2021, and 2022). Figures S1 and S2 show infant and neonatal mortality trends for each country.

**Figure 1.**
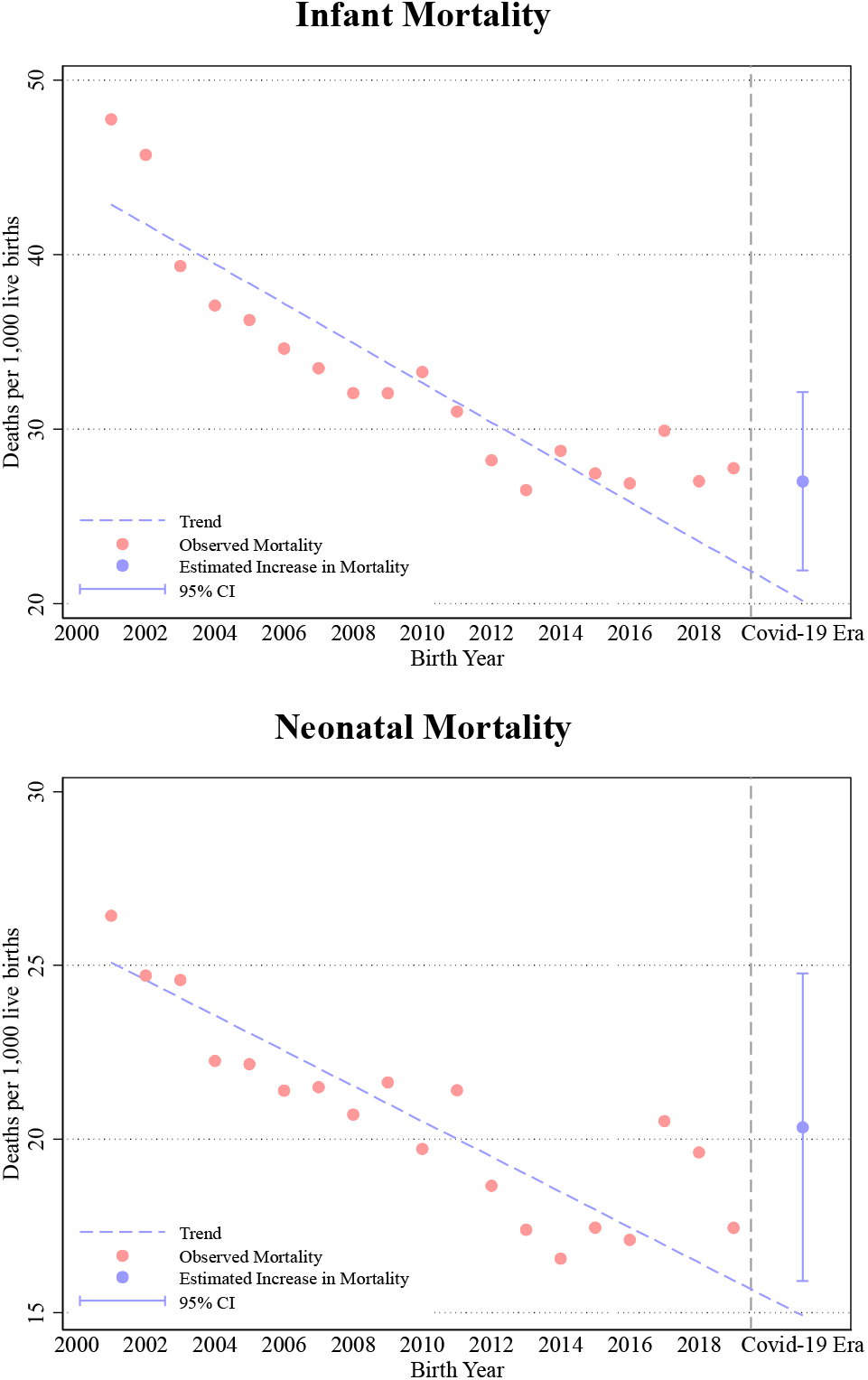
Infant and Neonatal Mortality by year. This figure shows infant (panel A) and neonatal (panel B) mortality rates from 2000-2022. Each point represents the number of infant or neonatal deaths per 1,000 live births in the respective year adjusted for cluster-fixed effects. The trend line was estimated using only data to the left of the dashed line. The Covid-19 era includes births from 2020, 2021, and 2022. The 95% confidence interval was estimated using a regression that included a yearly trend, an indicator for whether the birth occurred after 2020, cluster fixed-effects, and calendar month of birth fixed-effects. Years 1990 to 1999 were not included for presentation purposes but were included in the main analysis reported in Figure 2. Thus, this figure does not directly map to the interrupted time series regression estimates in Figure 2.

Our main interrupted time series estimates are plotted in Figure 2. In the pooled analysis, infant mortality was higher than expected by 9.9 deaths per 1,000 live births after March 2020 (95% CI 5.0, 15.0; p<0.01), a 22% increase above the expected rate of 44 deaths per 1,000 live births. Neonatal mortality was higher than expected by 6.7 deaths per 1,000 live births (95% CI 2.4, 11.1; p<0.01), a 27% increase above the expected rate of 25 deaths per 1,000 live births. Both types of mortality increased similarly across countries aside from Cambodia, where we observe a relatively large increase in infant mortality and a relatively small increase in neonatal mortality (Figure 2).

**Figure 2.**
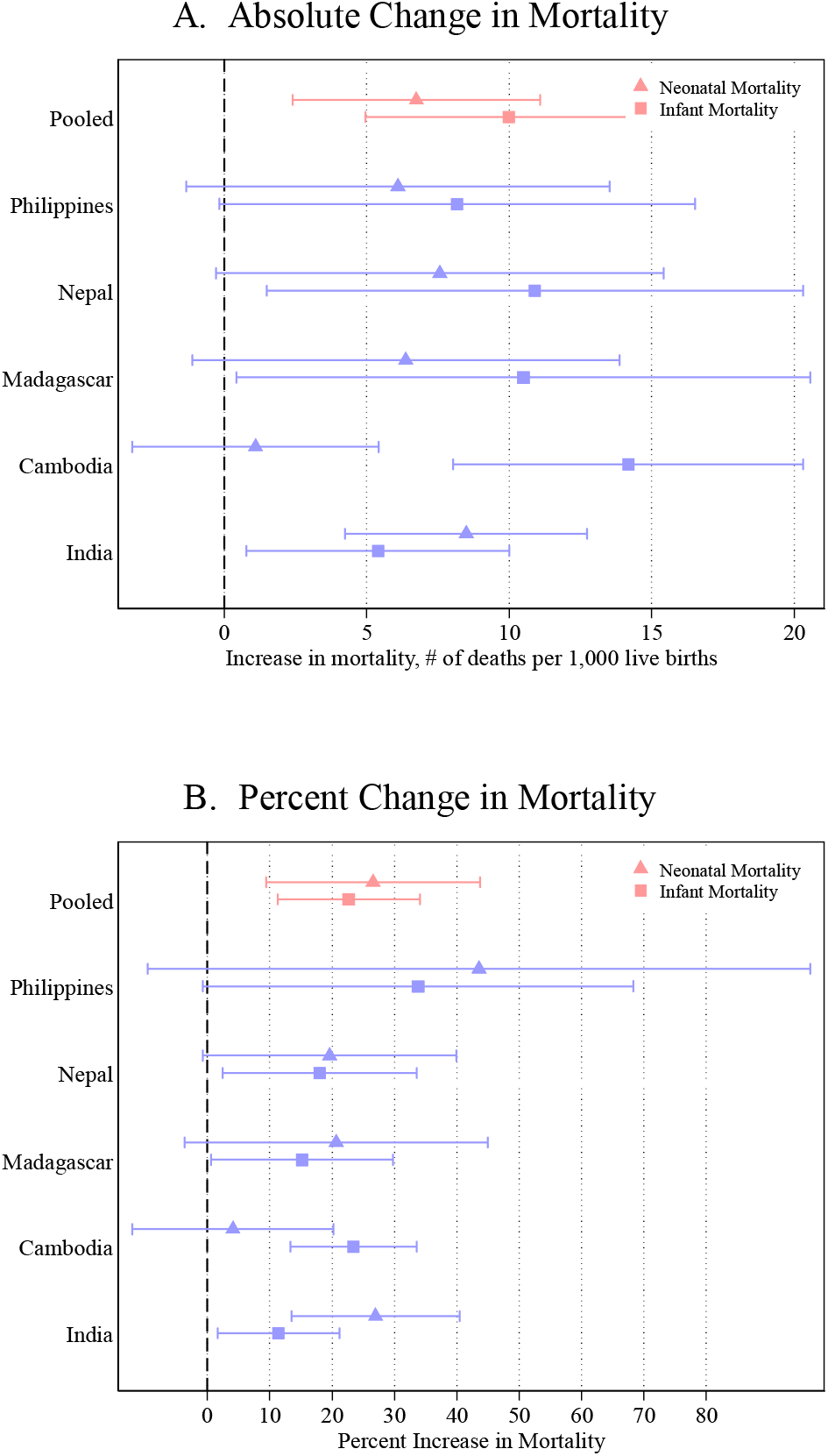
Change in infant and neonatal mortality following the start of the Covid-19 Pandemic. Estimates are from interrupted time series models described in equation 1 that include a monthly trend, an indicator for whether the birth occurred after March 2020, cluster fixed-effects, and calendar month of birth fixed-effects. The plotted points represent the increase in mortality relative to what was predicted by the pre-pandemic trend.

Figure 3 shows that the probability of receiving any antenatal care decreased by 3.8 percentage points relative to expectation (95% CI -4.9, -2.7; p<0.01) and facility deliveries decreased by 5.6 percentage points (95% CI -7.2, -4.0; p<0.01). Both maternal health care access indicators decreased in all countries aside from Madagascar.

**Figure 3.**
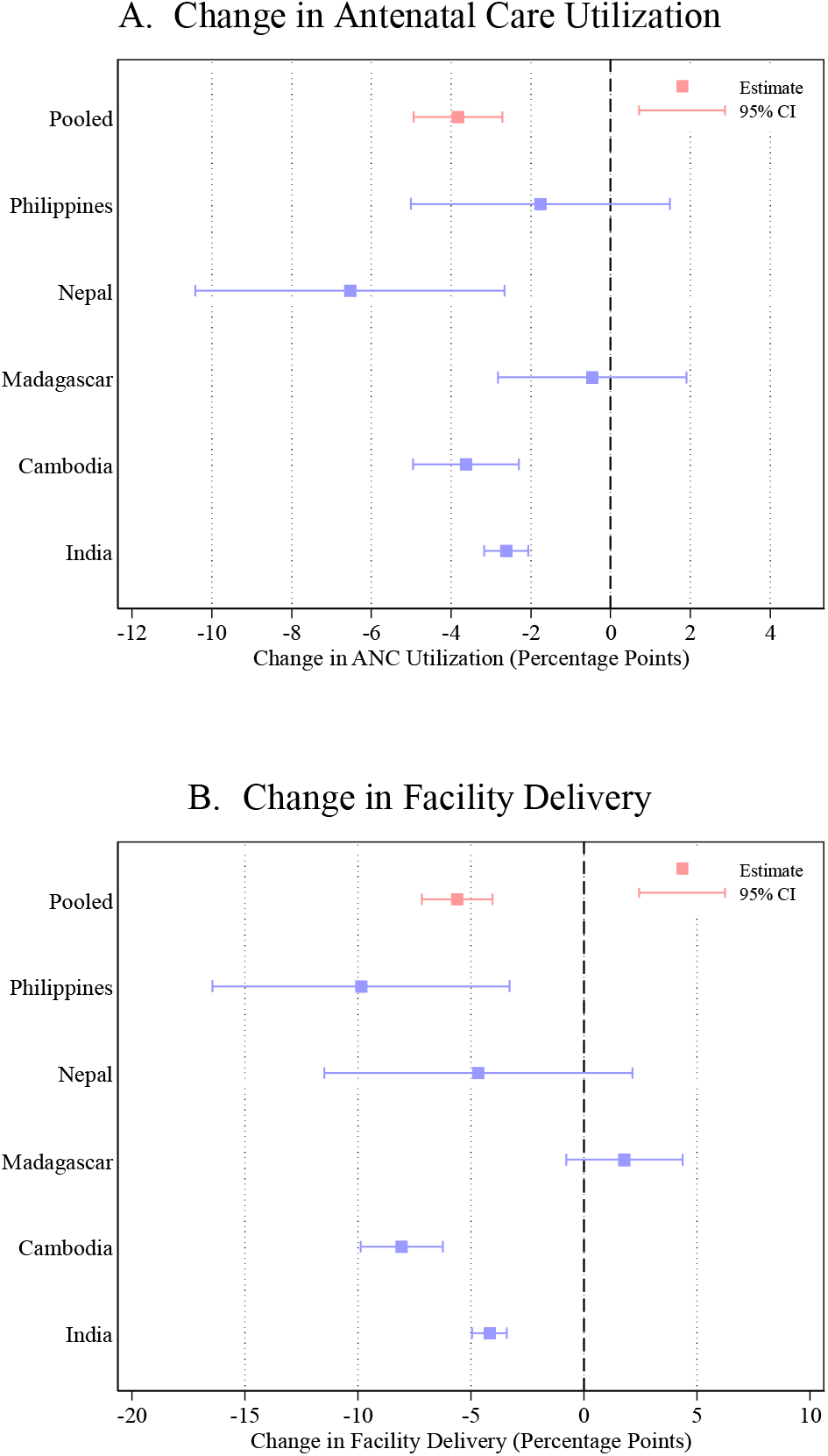
Change in infant and neonatal mortality following the start of the Covid-19 Pandemic. Estimates are from interrupted time series models that include a monthly trend, an indicator for whether the birth occurred after March 2020, cluster fixed-effects, and calendar month of birth fixed-effects. The plotted points represent the increase in mortality relative to what was predicted by the pre-pandemic trend.

In the sensitivity analysis of shortened baseline trends, we find positive and qualitatively similar increases in infant and neonatal mortality, with loss of statistical power after 2004 (Figure S3). Tables S3 and S4 show that infant and neonatal mortality regression estimates were robust to choice of regression weights and to exposure choices for children born between April 2019 and March 2020.

## Discussion

We observe large increases in neonatal and infant mortality in Cambodia, India, Madagascar, Nepal, and Philippines in the years following the start of the Covid-19 pandemic. These increases followed decades of declining infant and neonatal mortality and are equivalent to the mortality levels around 2012. In other words, the pandemic was associated with a loss of about a decade’s gains in infant and neonatal mortality. While the exact magnitude of the reversal in each country differed, and the uncertainty around the estimated effect sometimes crossed zero, every effect size was positive and ranged from 8% to nearly 40% increase above expected mortality rates.

In conjunction with the trend reversals in mortality, we also observe declines in facility births and use of antenatal care. While we cannot distinguish the extent to which the estimated mortality effects result from Covid-19 deaths or the disruptions to vital health services, these findings are consistent with consequences of disruptions to health care. Improvements in care before and during birth have been important drivers of reductions in neonatal and infant mortality (neonatal deaths make up more than half of infant deaths in our study countries). Inadequate conditions around birth (e.g. home birth) are important risk factors for neonatal deaths.^19^ While the large mortality increases are likely not explained completely by the modest reductions in antenatal care use and facility deliveries, our findings are suggestive of a decline in health care access, which could have limited use of other essential services as well. We also note that the fatality rate from Covid-19 infection is very low among infants and neonates.^20^

The extent to which the declines in the use of antenatal care and facility births was driven by supply-side factors (e.g. closure of facilities), demand-side factors (avoidance of healthcare facilities), or both, cannot be identified in this study. The experience in the rest of the world suggests that both demand-side and supply-side factors were involved in the observed changes in health care use. Here, we demonstrate that, in either case, indirect consequences of the pandemic were associated with reduced health care access and mortality increases.

Some limitations deserve further discussion. First, our study includes only 5 countries. Nationally-representative data in LMICs is sparse, and the patterns in other countries will emerge as more data is collected and released. Second, the global ubiquity of Covid-19 responses mean that we had to rely on interrupted time-series rather than another causal inference approach that uses a control group. The greatest challenge to inference is that we cannot establish a firm counterfactual beyond extrapolating past trends. While the reduction in infant and neonatal mortality has been consistent over time and appears linear, we cannot rule out the possibility that mortality started to increase prior to the start of the pandemic. Finally, having only 1-2 years of follow-up and limited sample size to assess effect sizes at different time points in the Covid-19 era means that we do not know how long the observed increases in mortality will take before resuming pre-pandemic trends.

In conclusion, we find large and consistent increases in neonatal and infant mortality in five LMICs following the start of the Covid-19 pandemic. These increases are equivalent to around a decade’s gains in mortality reductions. Helping to get reductions in neonatal and infant mortality back on track in LMICs should be a major global priority.

## Data Availability

All data produced in the present study are available upon reasonable request to the authors

