## Supplementary Appendix for "Infant and Neonatal Mortality During the Covid-19 Pandemic: An Interrupted Time Series Analysis From Five Low- and Middle-Income Countries"

Zachary Wagner^1,2^, Sam Heft-Neal^3^, Zetianyu Wang^1,2^, Renzhi Jing^4,5^, Eran Bendavid^4,5,6^

### **Supplementary Tables**

| Table S1. Regression results for infant mortality from figure 2 of main text | | | | | | |
| --- | --- | --- | --- | --- | --- | --- |
|  | (1) | (2) | (3) | (4) | (5) | (6) |
| VARIABLES | Pooled | India | Cambodia | Madagascar | Nepal | Philippines |
| Month (Trend) | -0.129*** | -0.109*** | -0.246*** | -0.172*** | -0.248*** | -0.034*** |
|  | (0.005) | (0.003) | (0.013) | (0.017) | (0.013) | (0.008) |
| Post March 2020 | 9.979*** | 5.393** | 14.175*** | 10.497** | 10.900** | 8.171* |
|  | (2.561) | (2.357) | (3.131) | (5.140) | (4.801) | (4.259) |
| Observations | 2,959,186 | 2,491,185 | 146,755 | 97,648 | 110,477 | 113,121 |
| Cluster Fixed-Effects | Yes | Yes | Yes | Yes | Yes | Yes |
| Month-of-year Fixed-Effects | Yes | Yes | Yes | Yes | Yes | Yes |
| Number of Clusters | 67754 | 58418 | 2942 | 1502 | 1657 | 3235 |
| Expected Mortality | 44.05 | 47.31 | 60.49 | 69.06 | 60.55 | 24.18 |
| Estimates are from interrupted time series models that include a monthly trend, an indicator for whether the birth occurred after March 2020, cluster fixed-effects, and calendar month of birth fixed-effects. Standard errors clustered by DHS cluster location are in parentheses. Expected mortality was estimated from recycled predictions using the Margins command in Stata. | | | | | | |

| Table S2. Regression results for neonatal mortality from figure 2 of main text | | | | | | |
| --- | --- | --- | --- | --- | --- | --- |
|  | (1) | (2) | (3) | (4) | (5) | (6) |
| VARIABLES | Pooled | India | Cambodia | Madagascar | Nepal | Philippines |
| Month (Trend) | -0.058*** | -0.060*** | -0.066*** | -0.042*** | -0.139*** | -0.007 |
|  | (0.004) | (0.002) | (0.008) | (0.012) | (0.010) | (0.007) |
| Post March 2020 | 6.743*** | 8.493*** | 1.098 | 6.371* | 7.565* | 6.099 |
|  | (2.218) | (2.168) | (2.207) | (3.825) | (4.007) | (3.793) |
| Observations | 2,959,186 | 2,491,185 | 146,755 | 97,648 | 110,477 | 113,121 |
| Cluster Fixed-Effects | Yes | Yes | Yes | Yes | Yes | Yes |
| Month-of-year Fixed-Effects | Yes | Yes | Yes | Yes | Yes | Yes |
| Number of Clusters | 67754 | 58418 | 2942 | 1502 | 1657 | 3235 |
| Expected Mortality | 25.37 | 31.48 | 26.81 | 30.82 | 38.58 | 13.99 |
| Estimates are from interrupted time series models that include a monthly trend, an indicator for whether the birth occurred after March 2020, cluster fixed-effects, and calendar month of birth fixed-effects. Standard errors clustered by DHS cluster location are in parentheses. Expected mortality was estimated from recycled predictions using the Margins command in Stata. | | | | | | |

| Table S3. Sensitivity analyses for infant mortality | | | | |
| --- | --- | --- | --- | --- |
|  | (1) | (2) | (3) | (4) |
| VARIABLES | Main | No Weights | Coding partial exposure as exposed | Dropping partially exposure |
| Month (Trend) | -0.129*** | -0.114*** | -0.131*** | -0.132*** |
|  | (0.005) | (0.002) | (0.005) | (0.005) |
| Post Pandemic Start | 9.979*** | 4.352*** | 9.569*** | 10.954*** |
|  | (2.561) | (1.275) | (2.063) | (2.608) |
| Observations | 2,959,186 | 2,959,186 | 2,959,186 | 2,919,103 |
| Cluster Fixed-Effects | Yes | Yes | Yes | Yes |
| Month-of-year Fixed-Effects | Yes | Yes | Yes | Yes |
| Number of Clusters | 67,754 | 67,754 | 67,754 | 677,52 |
| Expected Mortality | 44.05 | 49.58 | 43.93 | 44.29 |
| Estimates are from interrupted time series models that include a monthly trend, an indicator for whether the birth occurred after March 2020, cluster fixed-effects, and calendar month of birth fixed-effects. Standard errors clustered by DHS cluster location are in parentheses. Expected mortality was estimated from recycled predictions using the Margins command in Stata. Column 1 is the main estimate reported in the text. Column 2 estimates the same regression without using weights. Column 3 codes all children born between April 2019 and March 2020 as exposed to the pandemic. Column 4 drops all children born between April 2019 and March 2020 from the analysis. | | | | |

| Table S4. Sensitivity analyses for neonatal mortality | | | | |
| --- | --- | --- | --- | --- |
|  | (1) | (2) | (3) | (4) |
| VARIABLES | Main | No Weights | Coding partial exposure as exposed | Dropping partially exposure |
| Month (Trend) | -0.058*** | -0.057*** | -0.059*** | -0.059*** |
|  | (0.004) | (0.001) | (0.004) | (0.004) |
| Post Pandemic Start | 6.743*** | 5.745*** | 5.545*** | 7.248*** |
|  | (2.218) | (1.086) | (1.671) | (2.240) |
| Observations | 2,959,186 | 2,959,186 | 2,959,186 | 2,919,103 |
| Cluster Fixed-Effects | Yes | Yes | Yes | Yes |
| Month-of-year Fixed-Effects | Yes | Yes | Yes | Yes |
| Number of Clusters | 67754 | 67754 | 67754 | 67752 |
| Expected Mortality | 25.37 | 30.95 | 25.33 | 25.49 |
| Estimates are from interrupted time series models that include a monthly trend, an indicator for whether the birth occurred after March 2020, cluster fixed-effects, and calendar month of birth fixed-effects. Standard errors clustered by DHS cluster location are in parentheses. Expected mortality was estimated from recycled predictions using the Margins command in Stata. Column 1 is the main estimate reported in the text. Column 2 estimates the same regression without using weights. Column 3 codes all children born between April 2019 and March 2020 as exposed to the pandemic. Column 4 drops all children born between April 2019 and March 2020 from the analysis. | | | | |

| Table S5. Regression results for antenatal care utilization | | | | | | |
| --- | --- | --- | --- | --- | --- | --- |
|  | (1) | (2) | (3) | (4) | (5) | (6) |
| VARIABLES | Pooled | India | Cambodia | Madagascar | Nepal | Philippines |
| Month (Trend) | 0.001*** | 0.001*** | 0.001*** | -0.001** | 0.002*** | -0.000 |
|  | (0.000) | (0.000) | (0.000) | (0.000) | (0.000) | (0.000) |
| Post March 2020 | -0.025*** | -0.021*** | -0.024*** | -0.009 | -0.044*** | -0.008 |
|  | (0.004) | (0.002) | (0.004) | (0.010) | (0.008) | (0.007) |
| Observations | 451,190 | 364,347 | 28,965 | 21,360 | 19,879 | 16,639 |
| Cluster Fixed-Effects | Yes | Yes | Yes | Yes | Yes | Yes |
| Month-of-year Fixed-Effects | Yes | Yes | Yes | Yes | Yes | Yes |
| Number of Clusters | 65004 | 56046 | 2924 | 1496 | 1625 | 2913 |
| Expected ANC Use | 0.885 | 0.889 | 0.831 | 0.872 | 0.787 | 0.967 |
| Estimates are from interrupted time series models that include a monthly trend, an indicator for whether the birth occurred after March 2020, cluster fixed-effects, and calendar month of birth fixed-effects. Standard errors clustered by DHS cluster location are in parentheses. Expected mortality was estimated from recycled predictions using the Margins command in Stata. | | | | | | |

| Table S6. Regression results for facility delivery | | | | | | |
| --- | --- | --- | --- | --- | --- | --- |
|  | (1) | (2) | (3) | (4) | (5) | (6) |
| VARIABLES | Pooled | India | Cambodia | Madagascar | Nepal | Philippines |
| Month (Trend) | 0.002*** | 0.002*** | 0.003*** | -0.000 | 0.002*** | 0.002*** |
|  | (0.000) | (0.000) | (0.000) | (0.000) | (0.000) | (0.000) |
| Post March 2020 | -0.048*** | -0.036*** | -0.051*** | 0.011 | -0.044*** | -0.059*** |
|  | (0.006) | (0.004) | (0.007) | (0.011) | (0.015) | (0.013) |
| Observations | 597,868 | 488,996 | 36,989 | 24,668 | 25,817 | 21,398 |
| Cluster Fixed-Effects | Yes | Yes | Yes | Yes | Yes | Yes |
| Month-of-year Fixed-Effects | Yes | Yes | Yes | Yes | Yes | Yes |
| Number of Clusters | 65099 | 56380 | 2923 | 1234 | 1623 | 2939 |
| Expected Mortality | 0.548 | 0.839 | 0.556 | 0.370 | 0.340 | 0.640 |
| Estimates are from interrupted time series models that include a monthly trend, an indicator for whether the birth occurred after March 2020, cluster fixed-effects, and calendar month of birth fixed-effects. Standard errors clustered by DHS cluster location are in parentheses. Expected mortality was estimated from recycled predictions using the Margins command in Stata. | | | | | | |

### **Supplementary Figures**

| **Figure S1.** Infant mortality by year | |
| --- | --- |
| Pooled | India |
| 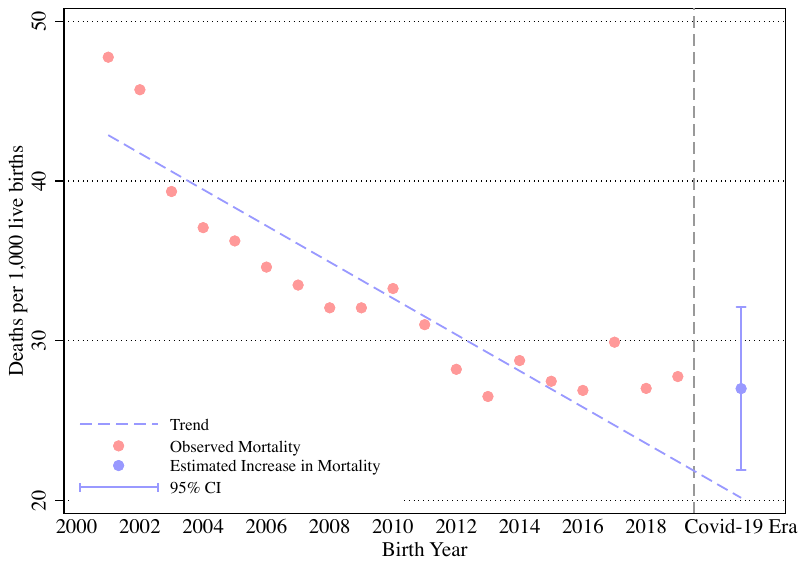 | 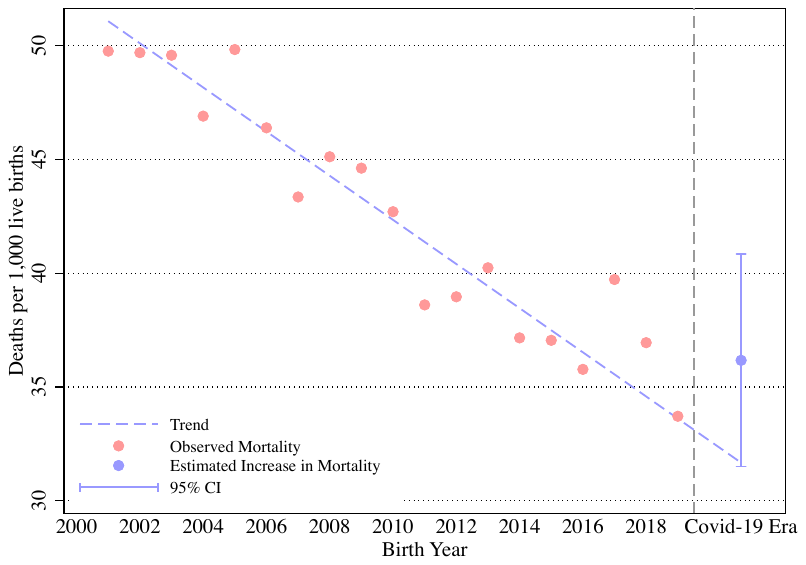 |
| Cambodia | Madagascar |
| 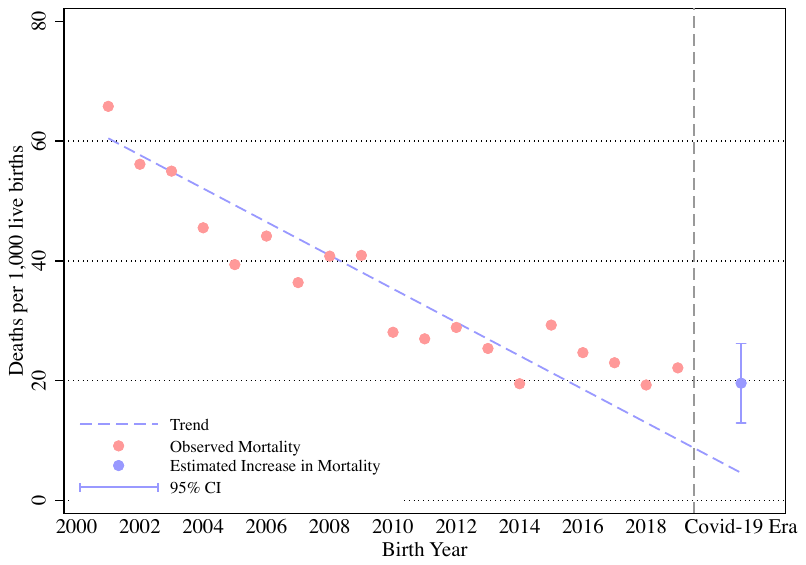 | 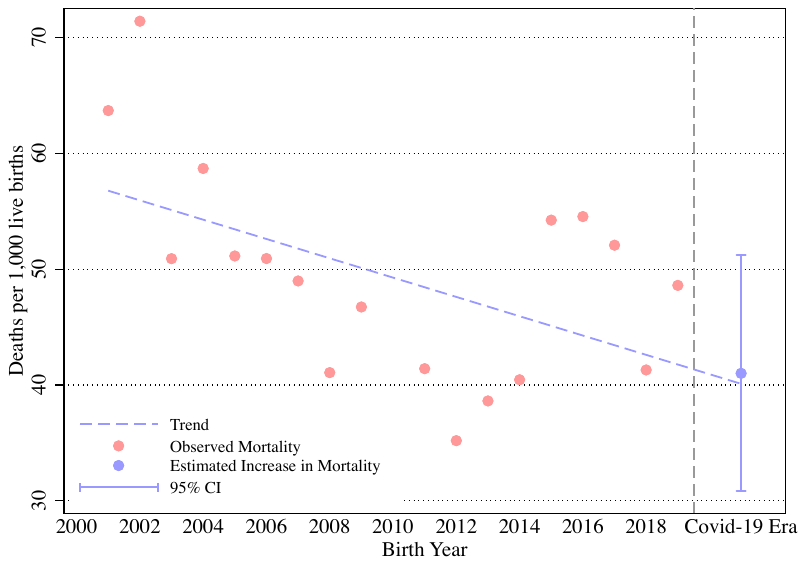 |
| Nepal | Philippines |
| 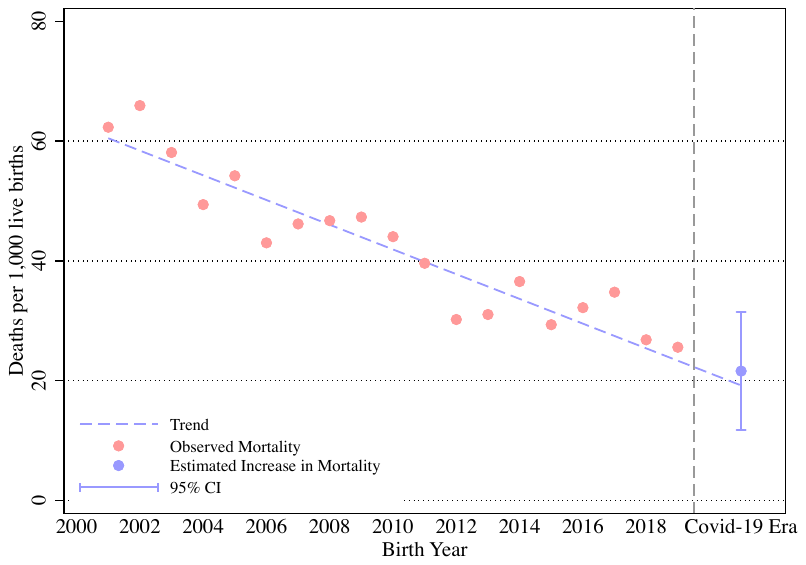 | 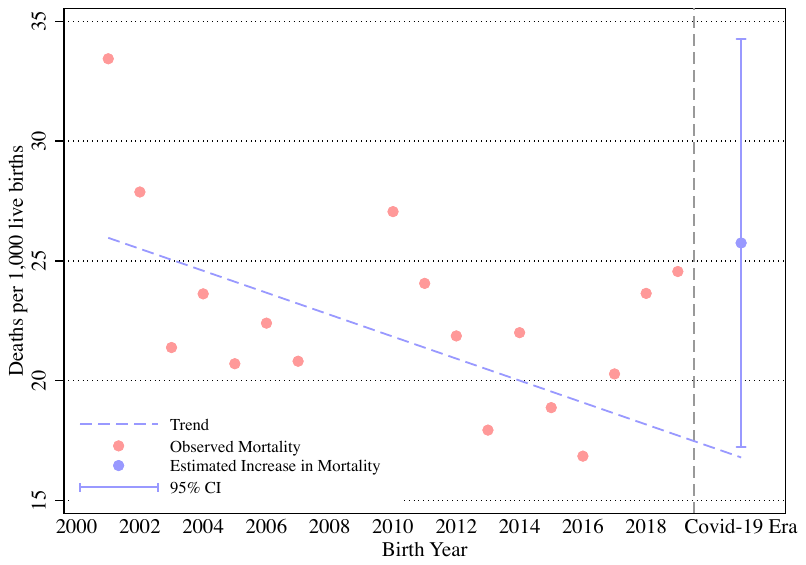 |
| Each point represents the number of infant or neonatal deaths per 1,000 live births in the respective year adjusted for cluster-fixed effects. The trend line was estimated using only data to the left of the dashed line. The Covid-19 era includes births from 2020, 2021, and 2022. The 95% confidence intervention was estimated using a regression that included a yearly trend, an indicator for whether the birth occurred after 2020, cluster fixed-effects, and calendar month of birth fixed-effects. Years 1990 to 1999 were not included for presentation purposes but were included in the main analysis. Thus, this figure does not directly map to the interrupted time series regression estimates in Figure 2. | |

| **Figure S2.** Neonatal mortality by year | |
| --- | --- |
| Pooled | India |
| 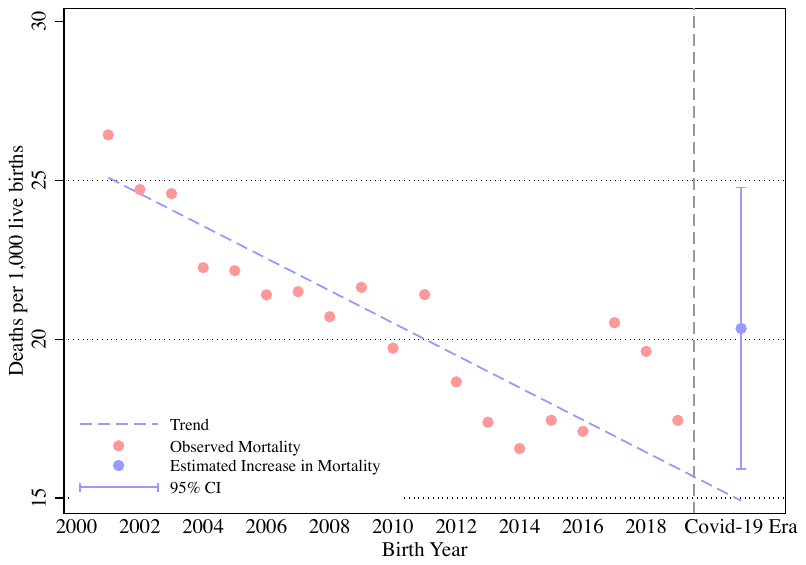 | 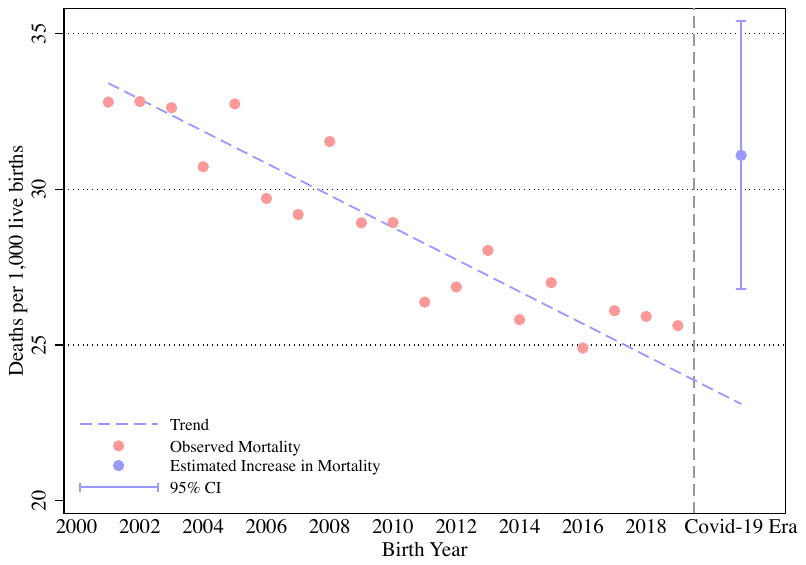 |
| Cambodia | Madagascar |
| 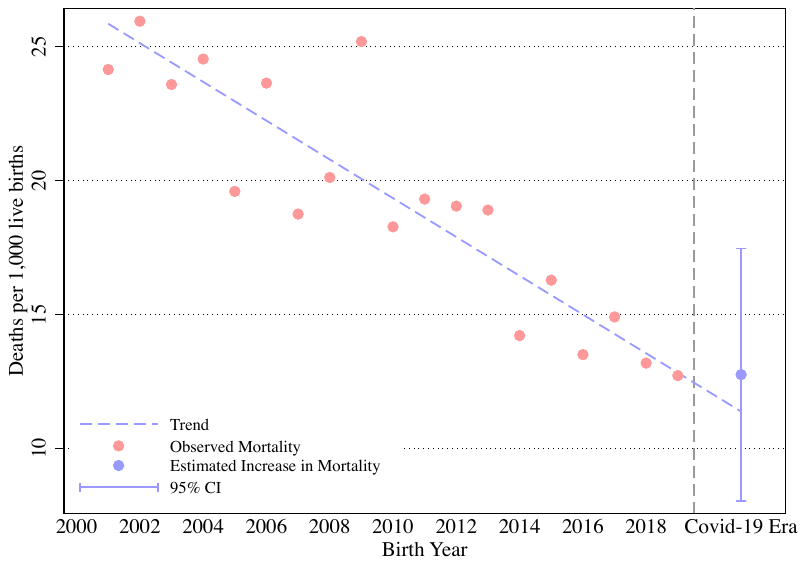 | 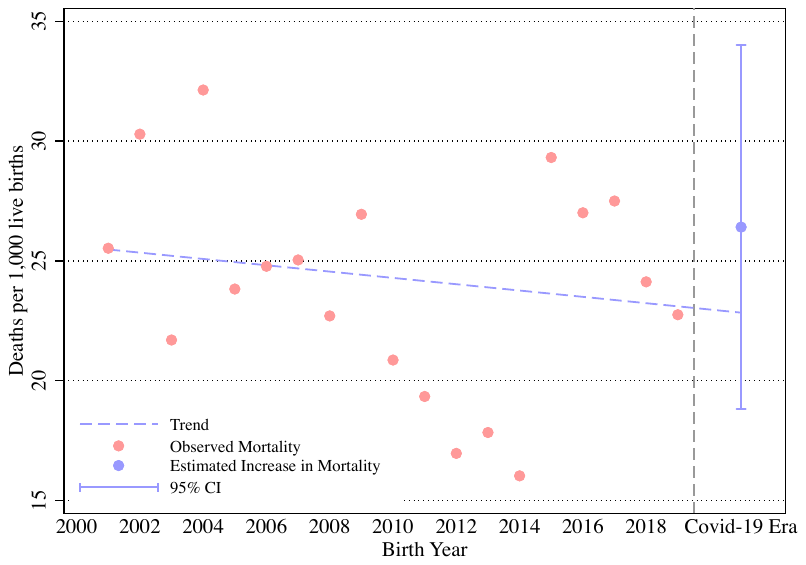 |
| Nepal | Philippines |
| 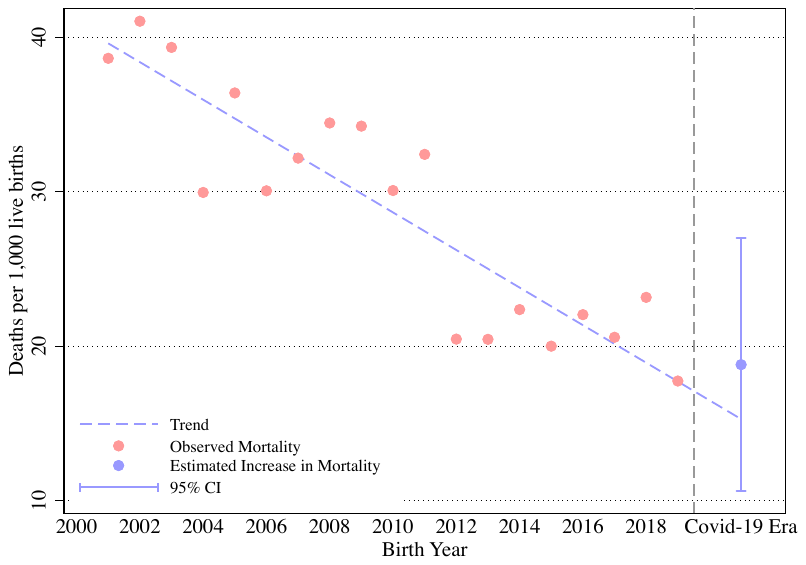 | 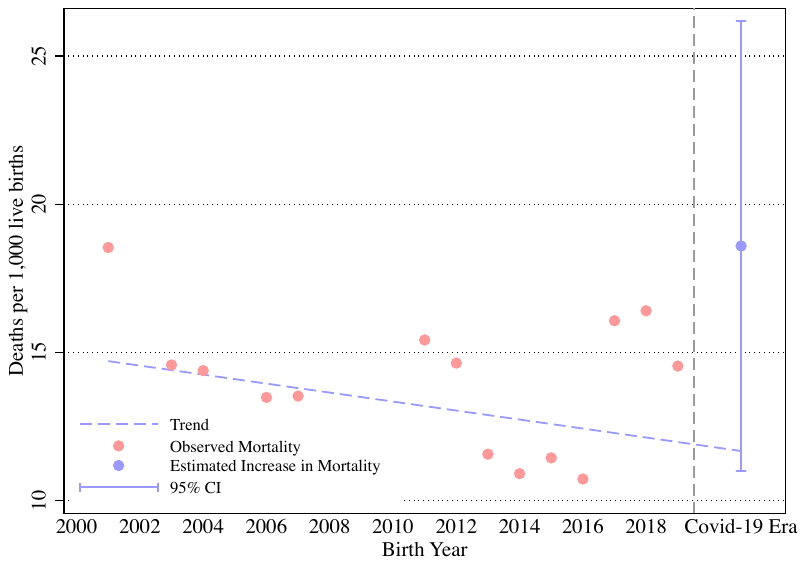 |
| Each point represents the number of infant or neonatal deaths per 1,000 live births in the respective year adjusted for cluster-fixed effects. The trend line was estimated using only data to the left of the dashed line. The Covid-19 era includes births from 2020, 2021, and 2022. The 95% confidence intervention was estimated using a regression that included a yearly trend, an indicator for whether the birth occurred after 2020, cluster fixed-effects, and calendar month of birth fixed-effects. Years 1990 to 1999 were not included for presentation purposes but were included in the main analysis. Thus, this figure does not directly map to the interrupted time series regression estimates in Figure 2. | |

#### Figure S3. Sensitivity in estimates of change in infant and neonatal mortality to different starting years

A. Infant Mortality Estimates

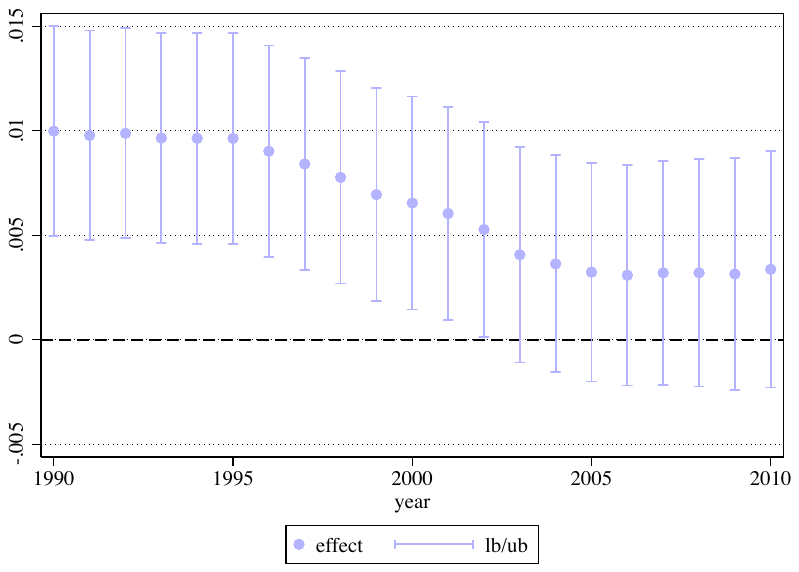

B. Neonatal Mortality Estimates

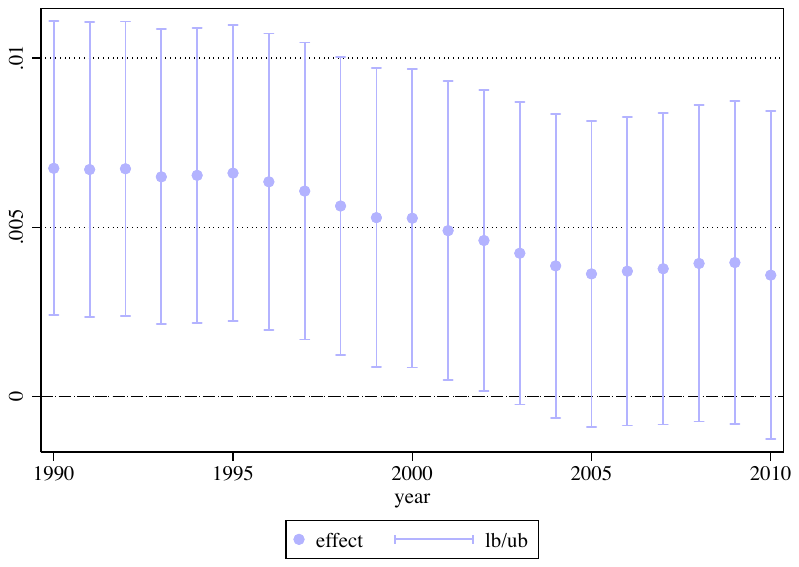

Estimates are from interrupted time series models that include a monthly trend, an indicator for whether the birth occurred after March 2020, cluster fixed-effects, and calendar month of birth fixed-effects. The plotted points represent the increase in mortality relative to what was predicted by the pre-pandemic trend.
